# Endogamy and high prevalence of deleterious mutations in India: evidence from strong founder events

**DOI:** 10.1101/2024.08.21.24312342

**Authors:** Pratheusa Machha, Amirtha Gopalan, Yamini Elangovan, Sarath Chandra Mouli Veeravalli, Divya Tej Sowpati, Kumarasamy Thangaraj

**Affiliations:** CSIR-Centre for Cellular and Molecular Biology, Hyderabad, Telangana - 500007, India; Academy of Scientific and Innovative Research (AcSIR), Ghaziabad - 201002, India; Department of Rheumatology and Clinical Immunology, Krishna Institute of Medical Sciences, Secunderabad, Telangana - 500003, India; Department of Rheumatology and Clinical Immunology, Nizam’s Institute of Medical Sciences, Hyderabad, Telangana - 500082, India; Department of Biotechnology, Bharathidasan University, Tiruchirapalli, Tamil Nadu - 620024, India

**Keywords:** Founder event, Endogamy, Novel variants, Runs of Homozygosity, Pharmacogenomics

## Abstract

Founder events influence recessive diseases in highly endogamous populations. Several Indian populations have experienced significant founder events and maintained strict endogamy. Genomic studies in Indian populations often lack in addressing clinical implications of these phenomena. We performed whole-exome sequencing of 281 individuals from four South Indian groups to evaluate population-specific disease causing mutations associated with founder events. Our study revealed a high inbreeding rate of 59% across the groups. We identified ∼29.2% of the variants to be exclusive to a single population and uncovered 1,284 novel exonic variants, underscoring the genetic underrepresentation of Indian populations. Among these, 23 predicted as deleterious were found in heterozygous state, suggesting they may be pathogenic in a homozygous state and are common in the endogamous groups. Approximately 40-68% of the identified pathogenic variants showed significantly higher occurrence rates. Pharmacogenomic analysis revealed distinct allele frequencies in CYP450 and non-CYP450 gene variants, highlighting heterogeneous drug responses and associated risks. We report a high prevalence of ankylosing spondylitis in Reddys, linked to *HLA-B*27:04* allele and strong founder effect. Our findings emphasize the need for expanded genomic research in understudied Indian populations to elucidate disease risk and medical profiles, eventually aiming towards precision medicine and mitigating disease burden.

## Introduction

India is a land of extraordinary human diversity in terms of cultural, social and religious practices, with over four thousand anthropologically well-defined population groups, who speak more than 300 different languages. Majority of the Indian populations practice strict endogamy, resulting in substantial barriers to gene flow. Consanguinity, i.e. union between close relatives is highly pervasive in Southern India^1^. The rich genetic diversity within these largely endogamous groups, underscores the need to increase the Indian ethnic representation in genetic data, crucial for advancing global precision medicine. Unfortunately, there exists a significant disparity in the inclusion of the non-European population in genomic studies. Capturing the genetic diversity of these underrepresented communities can address the existing disparities improving disease risk predictions, origin and early detection, diagnosis, clinical care, and precision medicine.

Founder events and population bottlenecks also shape the genetic constitution of Indian populations. Founder events occur when a new population is established by a small group of individuals separating from the ancestral population. This leads to reduced genetic diversity and distinctive allele frequency patterns, often resulting in a higher prevalence of rare recessive diseases^2^. Due to founder events, deleterious variations persist, more so when compounded by inbreeding practices. Such populations are valuable for examining how evolutionary processes influence disease genetics and evolution.

The small fraction of genetic variants that are not shared among populations, may also define their distinct metabolic phenotypes. Pharmacogenomics, which studies the impact of genetic makeup on drug response, is a promising avenue for personalized medicine. There are notable inter- and intra-ethnic differences in genetic variants associated with drug absorption, distribution, metabolism, and elimination (ADME)^3^. For example, inconsistent responses to 5-Fluorouracil among South Asian ethnic populations are attributable to DPYD gene variations^4^. However, most pharmacogenomic studies focus on European subjects, leading to a Eurocentric bias in drug dosing recommendations. Although limited studies exist profiling gene-drug interactions within India’s inter- and intra-ethnic diversity, the vast cultural diversity necessitates multiple frameworks.

In our previous study^5^, we identified 81 unique South Asian groups that exhibit stronger founder events than the Finns and Ashkenazi Jews, both known for a high incidence of recessive diseases linked to founder events. The strength of these founder events was quantified by analyzing the distribution of identity-by-descent (IBD) segments, which are stretches of genomic regions shared between individuals inherited from a common ancestor. While occasional reports mention specific rare diseases in select groups, comprehensive studies exploring the genetic composition and the potential for harboring deleterious variations predisposing these populations to certain diseases are lacking. In our present study, we performed an extensive interrogation of 281 high coverage whole exome sequences obtained from individuals belonging to four anthropologically distinct populations of India, each with a high IBD score, to (i) assess their levels of inbreeding (ii) identify the novel/population-specific and pathogenic variants (iii) estimate the distribution of pathogenic variants that might render populations-specific disorders and (iv) evaluate the genomic variants that affect drug metabolism in these groups. Further, in our findings, we reveal a widespread presence of Ankylosing Spondylitis (AS) in one of our study populations. We believe our study is one of the few that thoroughly explores the genetic landscape of Indian populations. This work marks an important step toward understanding Indian genetic diversity and has significant implications for the development of personalized medicine and public health strategies.

## Results

### Study populations and sequencing

Whole exome sequencing data was generated for all the collected samples from the four selected populations (Kalingas, Kallars, Reddys and Yadavs) of southern India, reported to have strong founder events^5^. We generated a final call set comprising of a total of 281 individual sequences across the four groups, inferred to be unrelated to the second degree. The analysis of the whole-exome data across the four populations demonstrated high-performance sequencing, achieving an average aligned read depth of approximately 99X in the target regions. The dataset was filtered using various parameters, including minimum read depth, proportion of missing data, and Phred quality scores, resulting in a high-confidence set of 140,921 variants across the four groups.

### Inbreeding coefficient reveals high endogamy

Inbreeding and consanguinity is a common practice in several ethnic groups in South Asia^6^. To understand the nature of inbreeding in our populations, we estimated the inbreeding coefficients for each individual and also the genealogical relationship between parents. Across populations, around 59% of the individuals had a positive inbreeding coefficient (56.1% in Kalinga, 87.5% in Kallar, 56% in Reddy and 38.4% in Yadavs). Mating type inference as depicted in Figure 1 showed that the Yadavs had comparatively more outbred individuals (61.6%), whereas the other three populations had a lower proportion of individuals whose parents are unrelated -43.8%, 12.5% and 44% for Kalinga, Kallar and Reddy, respectively (Table 1). We found no individuals exhibiting avuncular relationships between their parents.

**Fig. 1:**
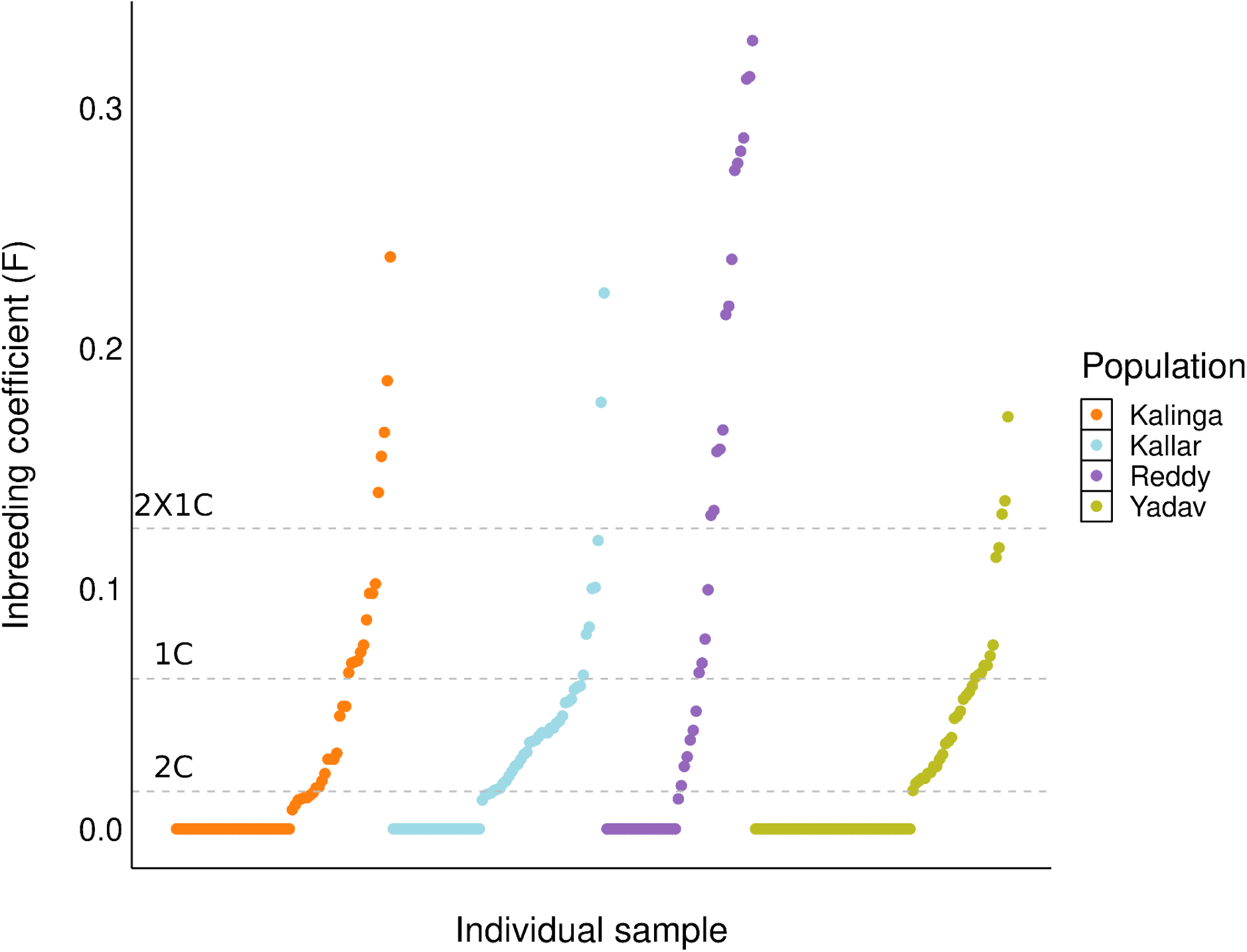
Proportion of inbred individuals across groups. The mating type inference for each individual was done. The gray lines indicate the inbreeding coefficient values for first cousins (1C), second cousins (2C) and double-first cousins (2x1C). A high level of inbreeding (∼59%) was detected across the four populations.

**Table 1.** Proportion of inbred and outbred individuals in the groups.

| Group | Total<br>samples | Outbred (%) | Inbred (%) |  |  |
| --- | --- | --- | --- | --- | --- |
|  |  |  | 1C | 2C | 1X2C |
| Kalinga | 73 | 43.8 | 13.7 | 31.5 | 11 |
| Kallar | 72 | 12.5 | 11.1 | 69.4 | 6.9 |
| Reddy | 50 | 44 | 6 | 18 | 32 |
| Yadav | 86 | 61.6 | 17.4 | 15.8 | 5.8 |

### Runs of homozygosity

Founder events, endogamy and consanguinity contribute to higher homozygosity in populations, with characteristic differences in length and number of homozygous tracts. Their sizes and numbers are informative; with the shorter ones reflecting autozygosity (IBD) and longer segments being suggestive of recent inbreeding events. We used runs of homozygosity (ROHs), *i.e*, contiguous genomic regions with identical maternal and paternal copies to elucidate the nature of inbreeding in the population groups. For our study, ROHs were grouped into five distinct classes - class A (1 - 2Mb), class B (2 - 4Mb), class C (4 - 8Mb), class D (8-16Mb) and class E (> 16 Mb) as depicted in Figure 2a (Supplementary Table 1). We detected ROHs in all 50 individuals from the Reddy group, 70 out of 73 from the Kalinga group, 71 out of 72 from the Kallar group, and 65 out of 86 from the Yadav group. Considering homozygous tracts exceeding 1.0 Mb in length revealed a wide range of values in the individuals, varying between a minimum of just 1 ROH tract to a maximum of 46 tracts through the genome. The ROH statistics across the groups are given in Table 2. The most extreme case was an individual from the Kalinga group, who had approximately 250.5 Mb of homozygous regions (Fig. 2b), covering about 7.8% of the autosomal genome, as can be seen in Figure 2c.

**Fig. 2:**
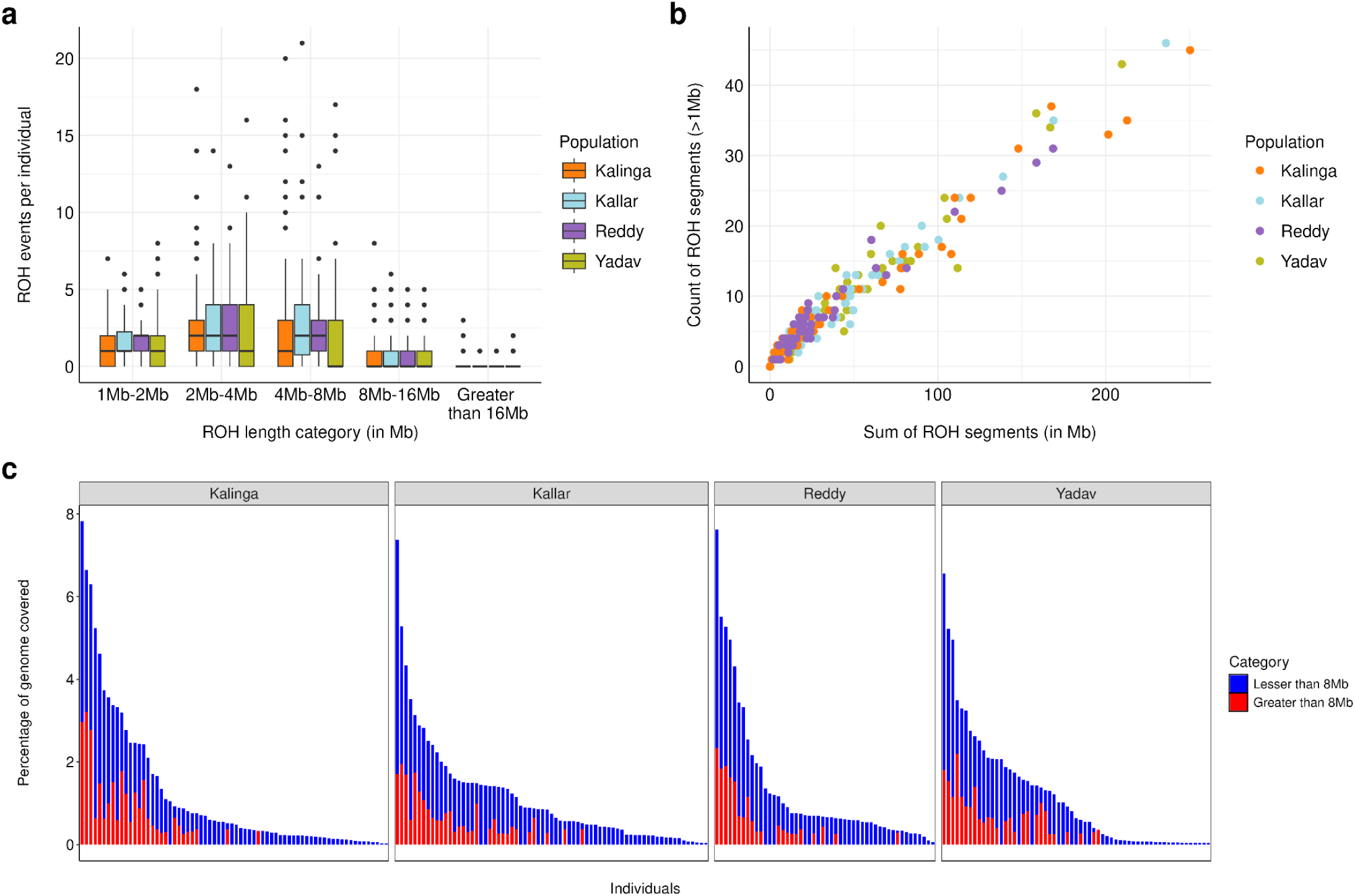
**a) ROH segments across the populations.** ROH regions spanning greater than 1Mb genomic region are considered to be significant and were distributed into five classes based on the length– class A (1-2Mb), class B (2-4Mb), class C (4-8Mb), class D (8-16Mb) and class E (>16Mb). **b) Relationship between the number and the sum of ROH segments per individual across the four population groups. c) Proportion of genome covered by ROH regions below 8Mb (1-8Mb) and above 8Mb.** A high proportion of genome occupied by ROHs in both categories indicates the presence of both founder events and recent inbreeding in the groups.

**Table 2.** ROH statistics across groups.

| Population | ROH statistics across groups |  |  |  |
| --- | --- | --- | --- | --- |
|  | Mean ROH<br>number per<br>individual | SD | Mean ROH<br>length per<br>individual | SD |
| Kalinga | 7.79 | 9.6 | 3.71 | 1.83 |
| Kallar | 8.86 | 8.15 | 6.23 | 2.84 |
| Reddy | 8.6 | 6.67 | 6.06 | 2.45 |
| Yadav | 6.52 | 9.39 | 4.12 | 3.74 |

Next, we attempted to see if the prevalence of ROH segments of a particular class correlates with the age of founder events. In all four groups, individuals exhibited larger proportions of homozygous tracts falling in class B and C (Fig. 2a). The occurrence of class C ROH segments indicates a shared ancestor approximately 12 generations ago. This observation partially coincides with the established timing of founder events documented for the groups in previous study^7^, wherein, the authors report the founder event to have taken place around 9 - 11 gBP (generations Before Present) in Kallars, 6 - 15 gBP in the Reddys and 4 - 7 gBP in the Yadav group. The Kalinga group was not a part of their study. While using a threshold of 20cM, we observed a major fraction (> 50%) of the homozygosity in each individual arising from the smaller segments, i.e < 8 cM. Also, the proportion of the autosomal genome occupied by the cumulative length of the smaller ROH tracts in the range of 1 - 8 Mb (class A - C) was higher than that occupied by longer tracts > 8.0 Mb (class D & E) for almost all the individuals in all four population groups, providing additional affirmation on the occurrence of founder events in these groups (Fig. 2c). The analysis also revealed that ∼ 38% of the samples display at least one ROH segment exceeding 8Mb. The equally higher prevalence of longer segments (> 8.0 Mb) aligns with the existing practice of endogamy and of consanguineous unions in the populations of South India. Both the founder events and inbreeding are observed to play a pivotal role in shaping the homozygosity in these groups.

### Inter-population comparison

Principal Component Analysis (PCA) was performed by combining genetic data of the 281 individuals with 1000 Genome phase 3 dataset (1kG_phase3). As expected, we observed that the groups aligned closely to the South Asian samples (Supplementary Fig. 1). In inter-population comparison, we found that around ∼ 29.2% of the total variants across all the four groups were found in just a single individual or population (population-specific), while 39.7% variants were shared by all the four populations (Fig. 3). At most, only 6.7% of variants are shared among three populations (Kalinga, Kallar, and Yadav), and 6.6% are shared between two populations (Kallar and Yadav). This highlights the distinct genetic constitution of the four populations.

**Fig. 3:**
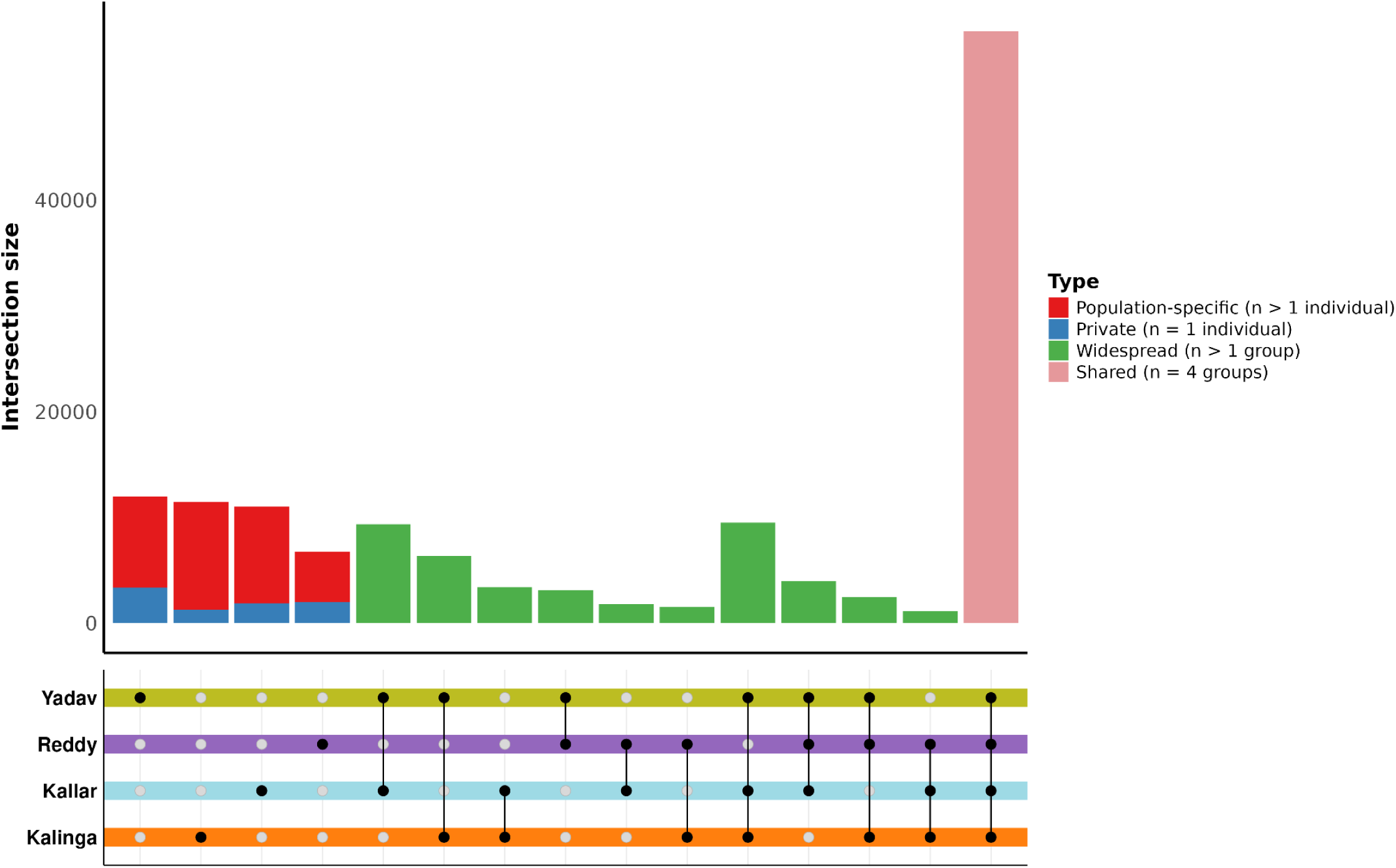
Upset plot showing the distribution of community-specific variants, and variants shared among the groups. Variants were classified as private (n=1 individual), population-specific (n>1 individual in a group), widespread (n>1 individual in more than one group) or shared (n>1 individual in all four groups). Around ∼ 29.2% of the total variants across all the four groups were found in just a single individual or population (population-specific), while 39.7% variants were shared by all the four populations.

### Novel variants

Post-filtering for each population separately, we detected a total of 73,599 high quality variants in Kalinga, 80,384 in Kallar, 79,255 in Reddy and 82,761 in Yadav (Supplementary Fig. 2a) in the exome data. Comparing these variants with nine widely used and publicly available population datasets (see methods), we segregated known and novel variants in each population (Supplementary Table 2). We identified 1,284 novel genetic variants, encompassing variants in the coding, UTR and splice regions, across the four groups. Population-wise, in Reddys, 526 novel variants (0.66% of the total variant set) were identified, while Kalingas had 472 exonic variants (0.64%) to be novel. Kallars presented 205 novel variants, constituting 0.26% of the total, followed by Yadavs with just 86 of them, accounting for 0.1% of the total (Supplementary Fig. 2b, Supplementary Fig. 3). Importantly, a comparison among different population groups revealed that the identified novel variants are exclusive to their respective group, with only a maximum of 4 novel variants being shared between the Yadav and the Kalinga groups, while only one novel variant is common among the Kalinga and the Reddys. Further, the asymmetrical distribution of these novel variants can be characterized as an enrichment of rare and missense variants, as the majority of the novel variants having a MAF (Minor Allele Frequency) between 2-5 and the number decreasing with increase in the MAF (Supplementary Table 2).

Given that we observe an excess of rare novel variants at functional sites in our populations, we consider the effect of these variants on fitness and selection using different approaches. After annotation with Annovar and Ensembl VEP against different databases, these variants were analyzed for their potential consequences (see methods). We detected a limited number of potentially deleterious novel variants - 7 in Kalingas, 4 in Kallars, 9 in Reddy and 3 in the Yadavs. These are either non-synonymous variants or are present in the splice sites. An important aspect to mark here is that none of the individuals display homozygosity for the novel potentially deleterious genetic variants (Table 3). We then predicted the impact of these variants on the stability of the protein. Of the 17 genes with missense variants, 15 of them displayed a destabilizing/unfavorable effect on the protein, while two of them in the *ACOT8* and the *MAP4K1* genes had a stabilizing/favorable effect.

**Table 3.**
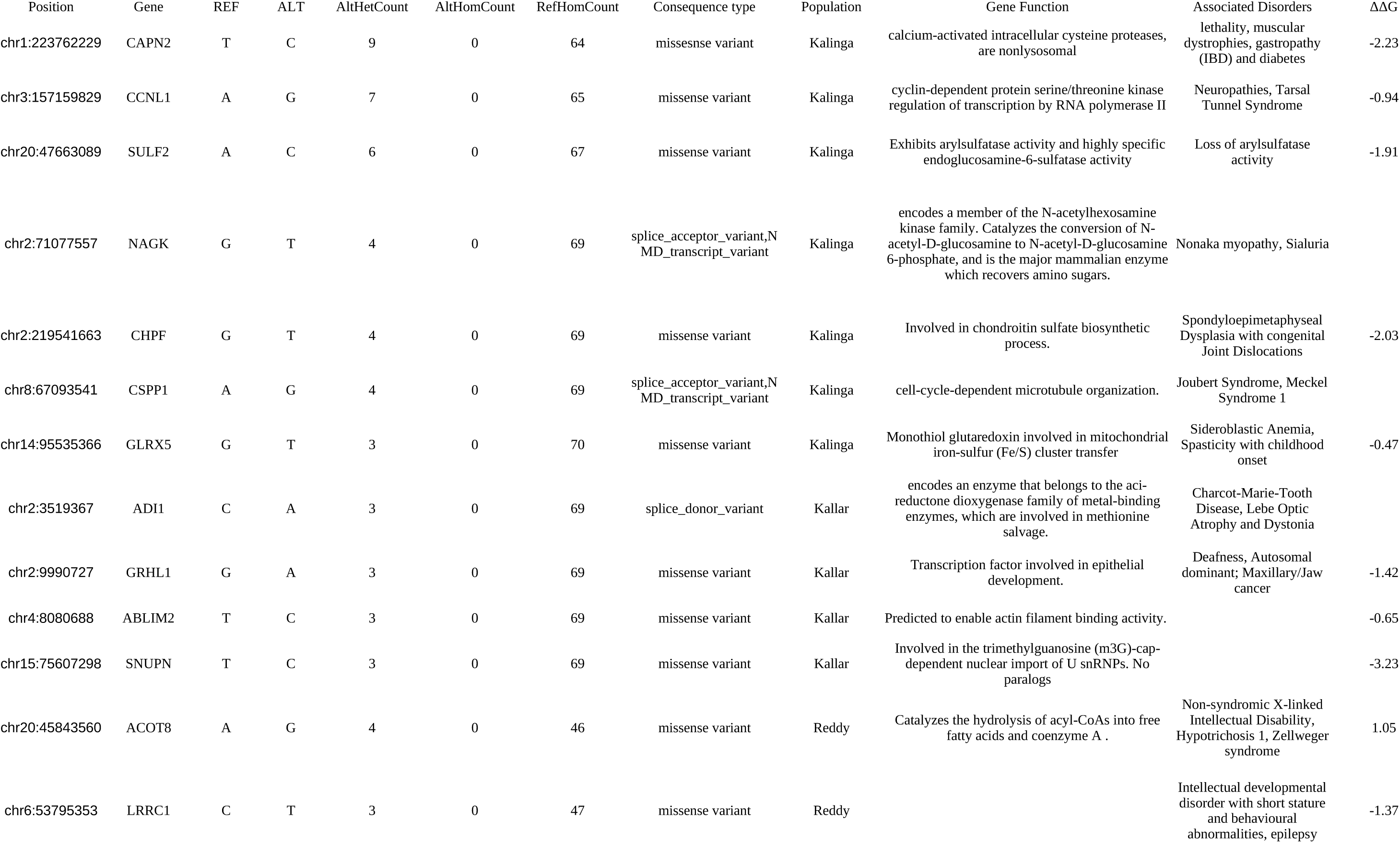
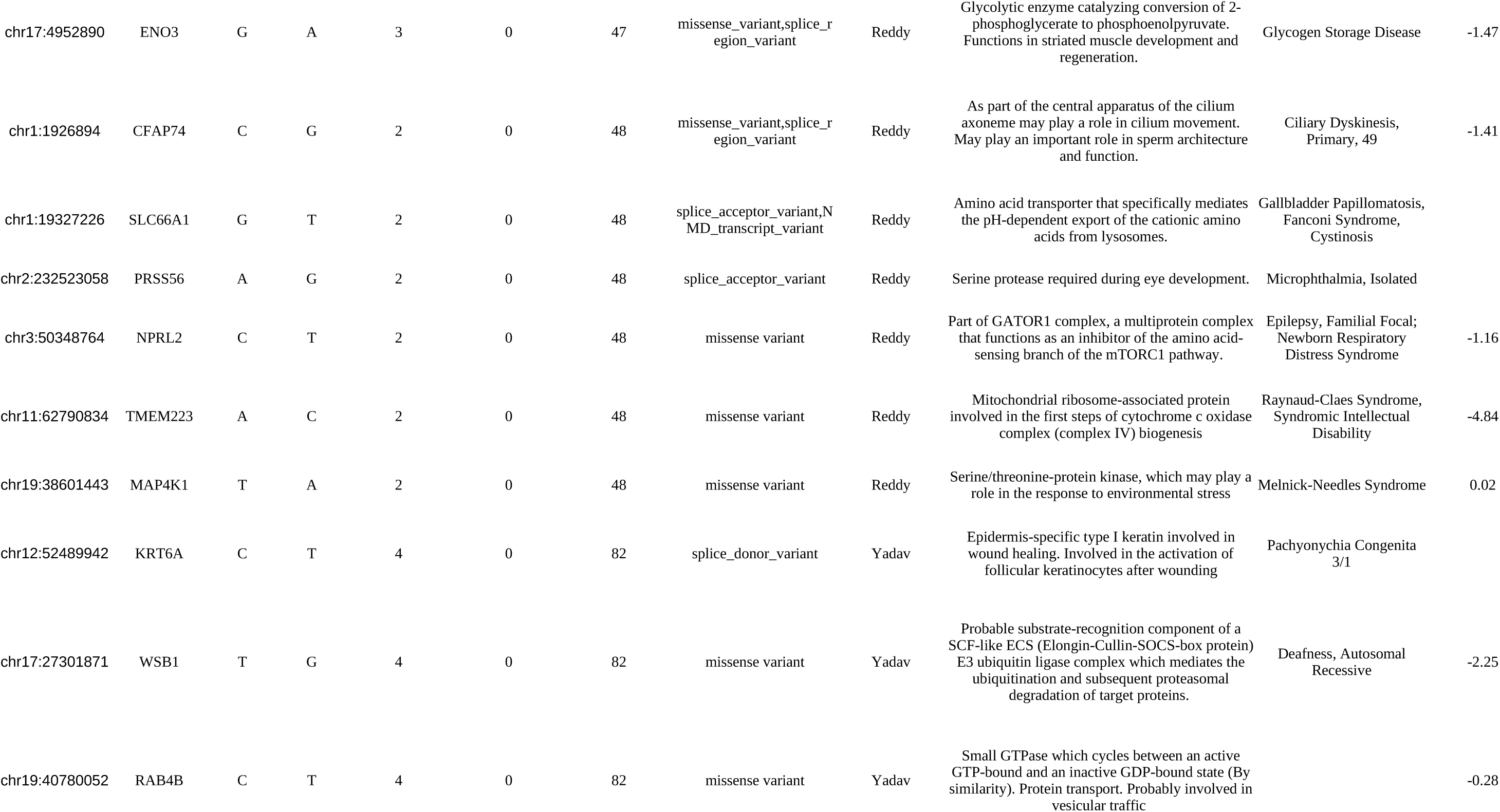
Novel variants predicted to be deleterious along with their impact on protein stability.

### Known variants and enrichment analysis

For the known variants that were annotated as pathogenic/likely pathogenic in ClinVar and thereby, referred to as “known deleterious” variants were majorly frameshift indels, stopgain or non-synonymous variants (Supplementary Fig. 4). We annotated 21 variants to be deleterious in Kallars, 30 each in the Kalingas and Reddys and 19 in the Yadavs (Supplementary Table 3). In comparison with the reported allele frequencies in the 1kG_phase3 dataset, a high proportion of these variants exhibited significantly (padj<0.05) higher AF, ranging between ∼33% in Kallars and Kalingas to ∼42% in Yadavs. Similar trends were also observed on comparisons with the GenomeAsia100k dataset (GAs100k) as depicted in Figure 4a. An inter-group comparison gave us six genes that are hosting pathogenic/likely pathogenic variants with a significant MAF in two or more groups (Supplementary Fig. 5). Similarly for the “known potentially deleterious” (predicted) variants, notable proportions were observed to be significantly (padj<0.05) enriched in the populations : ranging from approximately 39% in Yadavs to 47% in the Reddys, and around 65% in the Yadav and Kallar groups to 66% in the other two, on comparison with the AFs reported in the 1kG_phase3 and GAs100k datasets, respectively (Fig. 4b).

**Fig. 4:**
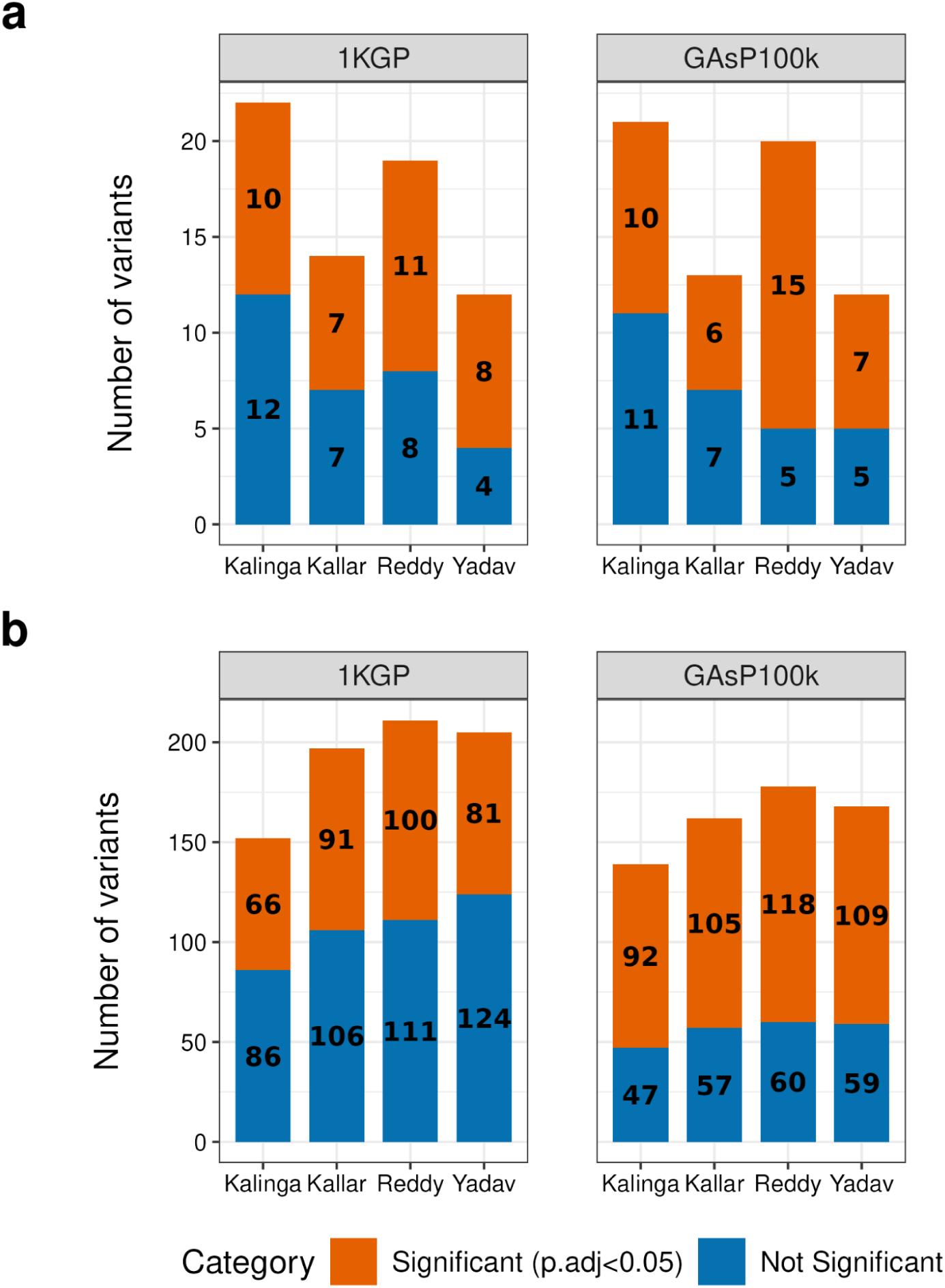
Significant known deleterious and known potentially deleterious variants across groups. The known variants categorized in the (a) deleterious and (b) potentially deleterious types across groups were assessed for the significance of the allele frequencies in comparison to those reported in 1000 Genome phase 3 dataset (left panel) and the GenomeAsia 100k dataset (right panel). Significance was determined using Fisher’s exact test followed by BH-correction for accuracy. The ones not reported in either of the datasets are omitted.

An over-representation analysis was performed to ascertain whether the high occurrence of founder variants in each group are enriched in specific pathways, molecular-functions, or cellular-components beyond what would be anticipated by random chance. The significant enriched terms (FDR<0.05) resulting from the Gene Ontology resource analysis in CPDB primarily included hydrolase activity, superoxide dismutase activity, t-UTP complex cellular component, along with others (Supplementary Table 4).

### Pharmacogenomic diversity

To delve into the pharmacogenomic profile of the study populations, we considered the pharmacogenetically important alleles listed in Tier1 of the Clinical Pharmacogenetics Implementation Consortium (CPIC) drug-gene pairs, along with the Pharmacogenomics KnowledgeBase (PharmGKB). The analysis was further classified into three different categories of genes - cytochrome-P450 (CYP450) genes, non-CYP450 genes and the Human Leukocyte Antigen (HLA) genes. We genotyped the first two groups of genes with the PyPGx^8^ tool. Herein, we observed substantial diversity within the CYP450 genes (Fig. 5a, 5b) - *CYP2B6*, *CYP2C19*, *CYP2C9* and *CYP2D6*. A carrier frequency exceeding 1% for several of the Tier1 pharmacovariants within these genes was observed across all the four groups (Table 4, Supplementary Table 5). However, for a majority of them, the AFs remain comparable to those documented for the South Asians, available on the CPIC website (Supplementary Table 6). For instance, *CYP2C9*3* had similar high allele frequencies as reported in the Indian populations^9^. The notable deviations were the *CYP2C19*3* having an AF of 11.11% and 15.34% in Kallars and Yadavs (compared to 2.73% in SAS), *CYP2D6*3* exhibited an AF of 13.7% in Kalingas and 20.45% in Yadavs (1.01% reported in SAS); and *CYP2D6*4* that showed an AF of 23.29% and 20.83% in Kalingas and Kallars, respectively (8.89% in SAS). Kallars had a representation of ∼6% UM along with ∼26% RM and ∼15% PM individuals for the *CYP2C19* gene that is involved in the metabolism of several xenobiotics, including some proton pump inhibitors and antiepileptic drugs. An occurrence of *CYP3A5*3* (A6986G) allele at an AF of ∼0.4, similar to the reported AF allele frequency outlined for South Indians^10^ was observed in the Kalingas. The variant has been assigned “No function” by CPIC for drugs like tacrolimus, eventually resulting in inter-individual differences - PM at ∼34%, IM at ∼51% and NM at ∼15%. The other three groups consist of samples with only NM and IM phenotypes.

**Fig. 5:**
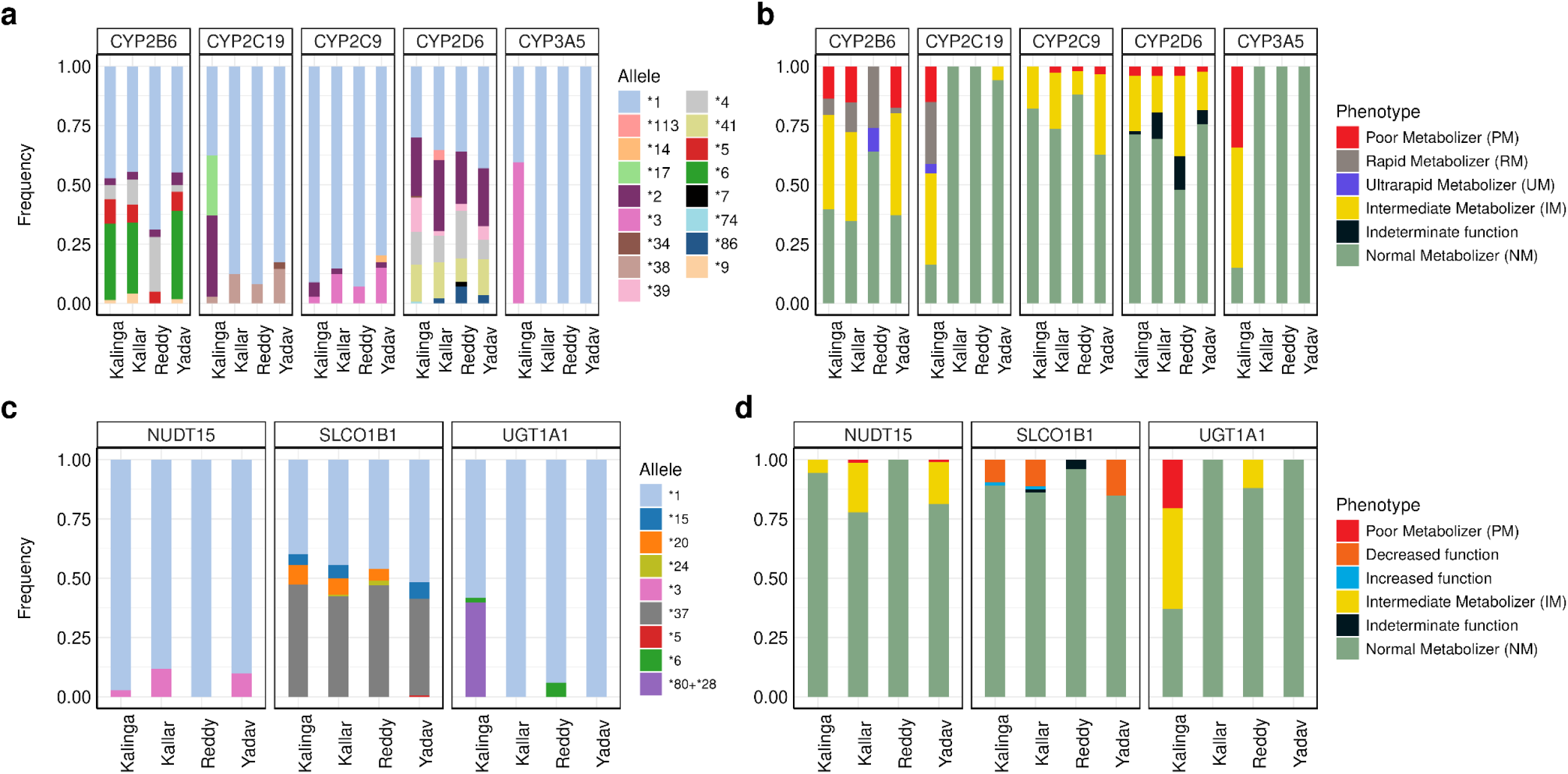
Genetic diversity and type of drug-metabolism for the PharmGKB VIP genes across groups. (a) and (b) focus on a few CYP450 genes, where they represent the allele frequency and the proportion of different types of metabolizers corresponding to the genotype of the individual, respectively. Similarly, (c) and (d) are for the non-cytochrome P450 genes. The diploid genotype of an individual determines the activity levels of the drug metabolizer protein.

**Table 4.** Details of VIP gene variants from PharmGKB across groups.

| Details of VIP gene variants from PharmGKB across groups |  |  |  |  |  |  |
| --- | --- | --- | --- | --- | --- | --- |
| Risk allele | Gene | Phenotype | No. of risk alleles in the genotyped individuals (%) |  |  |  |
|  |  |  | Kalinga | Kallar | Reddy | Yadav |
| rs1051266 | SLC19A1 | Arthritis, Rheumatoid | 69.18 | 70.14 | 53 | 59.3 |
| rs1801133 | MTHFR | Arthritis, Rheumatoid | 8.22 | 8.33 | 7 | 11.05 |
| rs3918290 | DPYD | PM | — | — | 3 | — |
| rs1801160 | DPYD | Neoplasms | 4.79 | 15.97 | 7 | 6.4 |
| rs1801265 | DPYD | Neoplasms | 17.12 | 27.78 | — | 27.91 |
| rs56038477 | DPYD | Neoplasms | — | — | — | 2.33 |
| rs2297595 | DPYD | Neoplasms | 6.85 | 4.17 | 11.46 | 13.37 |
| rs1801159 | DPYD | Neoplasms | 6.16 | 9.72 | — | 8.14 |
| rs2108622 | CYP4F2 | Increased warfarin dose requirement | 46.58 | 45.83 | — | 50.58 |
| rs2231142 | ABCG2 | Rosuvastatin toxicity | 17.81 | 12.5 | 10.2 | 7.56 |
| rs116855232 | NUDT15 | dosage; IBD, myelosuppression | 2.74 | 11.81 | — | 9.88 |
| rs2242480 | CYP3A4 | pain, postoperative | 41.78 | — | — | — |
| rs3745274 | CYP2B6 | HIV infections | 33.56 | 34.03 | — | 38.95 |
| rs4149056 | SLCO1B1 | Statin-related myopathy | 4.79 | 5.56 | — | 4.65 |
| rs9923231 | VKORC1 | Higher warfarin sensitivity | — | — | — | — |
| rs6025 | F5 | Thrombosis | — | 2.08 | — | — |
| CYP2B6*6 | CYP2B6 | Decreased function | 25.34 | 22.22 | — | 27.84 |
| CYP2B6*9 | CYP2B6 | Decreased function | 1.37 | 4.17 | — | 1.7 |
| CYP2B6*4 | CYP2B6 | Increased function | 6.16 | 10.42 | 18 | 2.84 |
| CYP2C9*3 | CYP2C9 | No function | 2.74 | 16.67 | 6 | 13.64 |
| CYP2C9*2 | CYP2C9 | Decreased function | 6.16 | 2.78 | — | 2.27 |
| CYP2C19*3 | CYP2C19 | No function | 2.74 | 11.11 | — | 15.34 |
| CYP2C19*2 | CYP2C19 | No function | 26.71 | — | — | — |
| CYP2C19*17 | CYP2C19 | Increased function | 23.29 | — | — | — |
| CYP2D6*3 | CYP2D6 | No function | 13.7 | 2.08 | 3 | 20.45 |
| CYP2D6*4 | CYP2D6 | No function | 23.29 | 20.83 | 25 | 5.11 |
| CYP2D6*41 | CYP2D6 | Decreased function | 13.7 | 13.19 | 10 | 12.5 |
| CYP3A5*3 | CYP3A5 | No function | 42.47 | — | — | — |
| NUDT15*3 | NUDT15 | No function | 2.74 | 11.11 | — | 9.09 |
| UGT1A1*6 | UGT1A1 | Decreased function, PM | 2.05 | — | 6 | — |
| HLA-B*15:02 | HLA-B | Increased risk of SCAR | 4.79 | 2.08 | 1 | 1.7 |
| HLA-B*57:01 | HLA-B | Increased risk of SCAR | 11.64 | 9.72 | 3 | 9.66 |
| HLA-B*58:01 | HLA-B | Increased risk of SCAR | 4.11 | 1.39 | 3 | 1.7 |

Secondly, for the non-CYP450 genes (Fig. 5c, 5d), we observed a high number of PM individuals for the drug atazanavir in only Kalingas. A total of 12 individuals (∼17%) who are homozygous for *UGT1A1*80+*28* are possibly at an increased risk for developing atazanavir- related hyperbilirubinemia^11^. We see a significant proportion of individuals with rs149056 variant in *SLCO1B1* gene, associated with statin-induced myopathy due to decreased efficacy in uptake of statins (mostly simvastatin and atorvastatin), in all groups, except the Reddys. For the identified Tier1 variants in our study groups, the majority exhibited an AF akin to those recorded for the South Asians. However, notable exceptions include rs2231142 in the *ABCG2* gene, associated with rosuvastatin pharmacokinetics^12^ demonstrating a significantly higher AF (FDR<0.05) in the Kalingas; rs3918290 and rs2297595 associated with DPYD deficiency-linked fluorouracil toxicity^13^ was found to be significantly enriched (FDR<0.05) in the Reddy and the Yadav groups, respectively. Additionally, rs2108622 (V433M) causing reduced *CYP42* activity and leading to the requirement of increased warfarin dosage^14^ displayed significance (FDR<0.05) in the Yadav group.

Thirdly, among the highly polymorphic HLA genes, we noted that Kallar, Kalinga and Yadav groups exhibited greater diversity in the *HLA-B* gene as compared to the *HLA-A* gene, while Reddys showed nearly similar proportion of allelic diversity for both the genes (as depicted in Supplementary Fig. 6). In general, we noted a considerable prevalence of approximately 31% of our study subjects carrying a *HLA-B* genotype associated with life-threatening drug toxicities including allopurinol-induced (*HLA-B*58:01*)^15^ or carbamazepine-induced (*HLA-B*15:02*) Stevens-Johnson syndrome/toxic epidermal necrolysis (SJS/TEN)^16^ and fatal hypersensitivity reactions to abacavir (*HLA-B*57:01*) in people with HIV infections^17^.

### Ankylosing Spondylitis (AS) in the Reddy population

We note a marked prevalence of the *HLA-B*27:04* genotype, accounting for 13% allele frequency within the Reddy samples. *HLA-B27*, a member of the major histocompatibility class I (MHC), is well established for its association with Ankylosing Spondylitis (AS) and in India, AS cases typically exhibit HLA-B27 positivity in the range of 80 - 90% ^18^. Through clinical collaboration with the Krishna Institute of Medical Sciences (KIMS), Hyderabad, Telangana, we successfully identified a significant proportion (54%, 7 out of 13 heterozygotes) of the genotype carriers testing positive for AS. We also observed that around ∼4.72% of diagnosed AS cases in the hospital records belong to the Reddy community, with a notable concentration of patients coming from the area of our sample collection and the adjacent villages. Furthermore, among the seven AS-positives from our dataset, two individuals have parents classifying as unrelated to each other, suggesting that besides consanguineous unions, founder events can also play a substantial role in certain diseases in Indian populations.

AS, a polygenic disorder, is one of the most common forms of inflammatory arthritis, predominantly affecting the axial skeleton and sometimes involving various other organs like the eyes, gastrointestinal tract; significantly diminishing the quality of life for those affected. Hence, we investigate the role of the second most common gene, *ERAP1* (Endoplasmic Reticulum Aminopeptidase 1), known to be associated with AS, in our sample set. We identified variants in the *ERAP1* gene and attempted to delineate the haplotypes in the AS-positive samples, focusing on eight widely studied variant sites. Alongside the wild type, we observed the prevalence of four different haplotypes - EPIMKDRE, EPIVKNQE, EPMMKDRE and ERIMKDRE (Table 5).

**Table 5.** List of ERAP1 allotypes in the AS-positive samples.

| Sample | rs3734016<br>(E56K) | rs26653<br>(R127P) | rs26618<br>(I276M) | rs2287987<br>(M349V) | rs30187<br>(K528R) | rs10050860<br>(D575N) | rs17482078<br>(R725Q) | rs27044<br>(Q730E) |
| --- | --- | --- | --- | --- | --- | --- | --- | --- |
| AS1 | E | P | I | V | K | N | Q | E |
| AS2 | E | P | M | M | K | D | R | E |
| AS3 | E | P | I | V | K | N | Q | E |
| AS4 | E | P | I | M | K | D | R | E |
| AS5 | E | R | I | M | K | D | R | E |
| AS6 | E | P | I | M | K | D | R | E |
| AS7 | E | P | M | M | K | D | R | E |

\*AS – Ankylosing Spondylitis , 1 -7 are the 7 positive samples

## Methods

### Study subjects and data generation

Objective of this study was to discern the genetic variants with potential implications in clinical phenotypes and diseases in populations with strong founder events. We selected four populations, which were reported to have high IBD scores and also have a huge census size of above or nearly 1 million individuals^5^. For the study, we collected samples of 101 individuals from the Kalinga population of Andhra Pradesh, 92 from the Kallars of Tamil Nadu, 66 from the Reddy group of Andhra Pradesh, and 94 from the Yadavs of Pondicherry (now Puducherry). To the best of our knowledge, all participants were in good health. Blood samples were collected from the volunteers and DNA was isolated using the phenol-chloroform method ^19^. Whole exome sequencing libraries were prepared using the TruSeq Exome library preparation kit and sequencing was performed on the Illumina Novaseq 6000. The paired-end sequencing was done to target a vertical coverage of 100X.

To have a whole genome representation for our study on clinically relevant variants, we genotyped a subset of 96 individuals (24 from each population group) for 700,604 variant sites on Infinium Global Screening Array-Multi Disease (GSA-MD v3.0) array. From each of the population, 24 random unrelated samples were selected and processed for GSA. This array was chosen for its relevant and curated clinical research markers from ClinVar and multi-ethnic exonic content from ExAC database or other published GWAS studies.

### Variant calling, filtering and quality metrics

Following basic quality control checks, adapter trimming was performed using Cutadapt v2.8. Alignment of the sequencing data to the human reference genome (build GRCh38) was performed using DRAGEN v3.9.3 to generate individual GVCF (genomic variant call format) files followed by joint genotyping to generate a single multi-sample VCF (variant call format) file for each population groups. An additional step of Variant recalibration was carried out using Genome Analysis Toolkit (GATK v4.4.0.0) by standard quality filters. We achieved a mean coverage of ∼99X on target regions for all the samples (Supplementary Table 8).

To exclude subjects with second degree relatedness, the kinship estimates were called using PLINK 2.0 - ‘KING’ and removed one sample from each pair having a kinship value > 0.0884. Further, low quality samples with less than 50X coverage and more than 50% missingness were removed. Post filtering of the samples, we have a final call set consisting of a total of 281 individual sequences (Kalinga-73, Kallar-72, Reddy-50 and Yadav-86) healthy individuals, inferred to be unrelated to the second degree. Stringent variant filtering strategies were applied for each of the groups separately to produce a high quality dataset. We used VCFtools v0.1.16 to exclude variants that were (i) non-biallelic (ii) genotyped in less than 95% of the samples (iii) Genotype Quality (GQ) less than 30 (iv) minimum read depth (minDP) less than 8 (v) presented a Hardy Weinberg test value p < 10^-6^ and (vi) Minor Allele Frequency (MAF) less than 0.02.

Post filtering, to assess the probability of false positives, the transition-transversion (ts/tv) and the concordance rate with dbSNP was estimated. The values are in accordance with the expected for exome studies. Further, to validate that the selected groups are non-overlapping, we computed the Weir and Cockerham F_st_ estimates between the groups (Supplementary Table 8). We considered pairs with weighted F_st_ < 0.004 to be overlapping^5^. However, we do not find any overlap between the groups suggesting that they do not intermarry. Analyzing the exome data for each population separately, we detected a total of 73,599 high quality variants in Kalinga, 80,384 in Kallar, 79,255 in Reddy and 82,761 in Yadav (Supplementary Fig. 1a).

For the GSA dataset, we conducted sample filtering to eliminate samples with a missingness exceeding 2% and those with any sex ambiguity. However, this step did not result in any sample loss. In the subsequent variant quality filtering steps, we removed duplicate variant sites and variants with less than 95% genotype rate across all samples. The genotyping rate for the final GSA dataset was ∼0.99 (Supplementary Table 7).

### Variant annotation

The post-QC exome dataset as well as the GSA data for each population was annotated both by Ensembl Variant Effect Predictor (VEP v102) and Annovar.

### Inbreeding and mapping regions of homozygosity

To detect the inbred individuals and infer the parental mating type, we used the FSuite v.1.0.4 pipeline^20^. It involves a method of estimating the individual inbreeding coefficients, obtained as F-median. We used the filtered exome files to generate 100 (default) random submaps. The proportions of the outbred and inbred - first cousins, second cousins and double-first cousins were calculated for each group.

The ROH segments were called from the exome dataset pruned for LD using PLINK; sliding window = 50kb, step size = 5 Single Nucleotide Polymorphisms (SNPs) and r^2^ threshold = 0.5. The homozygous regions were called for each sample with PLINK v1.9, the parameters optimized for WES^21^. This involves a sliding window of 50kb without any heterozygous sites, while keeping the rest of plink parameters at default (plink options : --homozyg-snp 50,-- homozyg-window-het 0). Only ROH with a length of 1Mb were selected. The ROH segments were binned into five-length classes : 1-2Mb, 2-4Mb, 4-8Mb, 8-16Mb and >16Mb, identified as class A-E, respectively. Each ROH length class represents the estimated number of generations from a common ancestor, calculated as 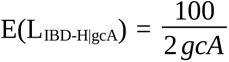, where E(LIBD-H|gcA) is the length of the ROH segment and gcA stands for the number of generations from the common ancestor^22^. Based on the assumption that 1cM equals 1Mb, we could estimate that the ROH classes from class A to class E, date back to approximately 50, 20, 12.5, 6 and 3 generations ago. The number of ROH segments, total length of these segments and the mean ROH length for each individual class was calculated.

### Population variant analysis

The final filtered variant call set for each of the population groups was categorized into rare (MAF 2-5), common (MAF 5-25) and very common (MAF >25) (Supplementary Table 2). Due to small sample size for the groups, we had filtered out the variants with MAF < 2 to achieve a high-confidence variant set.

To identify novel variants, we compared the identified and filtered variants from each group against nine known datasets namely - 1kG_phase3^23^, GAs100k^24^, gnomAD exome dataset, dbSNP151, the draft human pangenome^25^, ClinVar v.20221231, TopMed freeze 8 (https://bravo.sph.umich.edu/freeze8/hg38/downloads), and two Indian-specific datasets - one of the Andamanese^26^ and the second a collection of 836 refined Indian clinical research exome dataset^27^. To check if a variant is shared in a database, we matched the chromosome, position and both the REF and ALT alleles. The shared variants are referred to as the Known variants and the rest are called the Private variants with respect to the dataset of comparison. Finally, we made a final collection of variants that are totally unique to the populations in study and call them the Novel variants.

### Characterization and functional enrichment of the known and novel variants

Novel potentially deleterious variants - We put two different criteria to classify any variant to be putatively deleterious. One, from the VEP annotated vcf files, we filtered for the variants that were deemed to be of High Impact by VEP (such as frameshift, splice-site, stop-gain) and High Confidence (HC) by the Loss of Function (LoF) plugin. Two, we looked separately for *in silico* predictions by SIFT^28^, PolyPhen2^29^, and CADD v1.6^30^. Any variant annotated by the first two to be “deleterious” and “probably_damaging”, respectively; and having a CADD score above 30 (i.e the variant lies in the top 0.1 of deleterious variants in the human genome) were taken into consideration. Finally, the variants that pass either of our two criteria were classified to be potentially deleterious, in this case Novel potentially deleterious variants.

Known deleterious variants - These are the known variants for each population group that are annotated as “pathogenic/likely pathogenic” by ClinVar in the Annovar annotated files.

Known potentially deleterious variants - The known variants underwent a similar filtering strategy as for the novel potentially deleterious variants. Additionally under our second criterion, along with SIFT, PolyPhen and CADD, the known genomic variants were also annotated by two other *in silico* tools and those confirmed to be “deleterious” by LRT ^31^ and “disease causing” by MutationTaster^32^; were considered to be potentially deleterious.

For the known deleterious and potentially deleterious variants, the AF (allele frequency) was compared against 1kG_phase3 and GAs100k datasets for significance by performing Fisher’s exact test followed by Benjamini-Hochberg correction for FDR (False Discovery Rate) to account for multiple testing. The ones with p.adj < 0.05 are considered to be significant.

An over-representation analysis for the biological pathways of the genes mapped by the known deleterious/potentially deleterious variants was performed in the ConsensusPathDB program (http://cpdb.molgen.mpg.de/). As sensitivity analysis, we are calling a known variant to be a founder variant only if present in a significantly higher allele frequency (padj < 0.05) in the population (both compared to 1kG_phase3 and GAs100k dataset). The genes with significant known/potentially deleterious variants were given as input to the ConsensusPathDB (CPDB) program. The selected genes were the target and the complete list of genes present in the exome target sites was uploaded as reference, while selecting the GO pathway option. We incorporated a significance criteria of FDR < 0.05 in selecting the GO terms.

### Effect of novel potentially deleterious variants on protein stability

Following the *in silico* prediction of novel variants, we proceeded to assess the potential impact of these predicted novel deleterious variants on protein stability. This was done with the help of I-Mutant 2.0^33^, a Support Vector Machine (SVM) based tool that predicts the impact of single point mutations on the protein stability, given the protein structure or sequence. For each missense novel deleterious variant, we determined whether it exhibited a destabilizing effect (negative ΔΔG) or a stabilizing effect (positive ΔΔG) on the protein under standard conditions of pH 7 and room temperature. Here, ΔΔG is the predicted free energy difference and expressed as: ΔΔG= ΔGmutant − ΔGwild-type.

### Pharmacogenetics and *ERAP1* haplotype estimation

Pharmacogenomic variants associated with specific gene-drug responses were mined from the PharmGKB database at https://www.pharmgkb.org/ with clinical annotation of level 1A or 1B, since these are the variants demonstrating the highest evidence for actionable clinical implications of gene-drug associations. Allelic and the gene phenotype interpretations for the star allele genes (both cyt P450 and non-cyt P450 genes) was done using PyPGx v0.20.0^8^ tool. Using the genomic data, the PyPGx tool predicts the PGx genotypes and phenotypes, namely Normal Metabolizer (NM), Intermediate Metabolizer (IM), Poor Metabolizer (PM), Rapid Metabolizer (RM) and Ultrarapid Metabolizer (UM). For the HLA-A and HLA-B genes, xHLA v.1.2 ^34^ tool was used which utilizes the aligned bam files as input. Further, we computed the allele frequencies for the actionable genetic variants and their corresponding phenotypes by analyzing the output from the above two tools.

To compute the protein haplotypes of *ERAP1* gene, we constructed the haplotypes as a combination of eight amino acids at the coding SNP positions - rs3734016 (E56K), rs26653 (R127P), rs26618 (I276M), rs2287987 (M349V), rs30187 (K528R), rs10050860 (D575N), rs17482078 (R725Q) and rs27044 (Q730E), the haplotype represented in the same order as the sites mentioned here. These sites were selected as they are the most commonly reported variant sites in the gene and are also the ones mostly studied in connection with AS.

## Discussion

Founder events and population bottlenecks significantly influence genetic diversity and disease risk profiles in contemporary populations. These events cause allelic frequency shifts that have resulted in prominent genetic discoveries, notably in the Ashkenazi, Finnish, Amish, Icelandic and others. Endogamy/consanguinity contribute to increased homozygosity beyond what random mating would predict. Higher incidences of recessive disorders are typically found in founder as well as endogamous communities. In our previous study on South Asian populations^5^, we identified 81 out of 263 South Asian groups that experienced founder events stronger than the archetypal Finnish and Ashkenazi Jewish populations. India is characterized by diverse populations practicing strict endogamy. However, comprehensive studies on the disease risks and pharmacogenomic profiles of the Indian populations are scant.

Our current study reports high levels of endogamy in four Indian populations previously identified as having strong founder events. We depict a high percentage of ∼59% of the total individuals to be inbred across the four groups, and present both recent and ancestral parental relatedness as compelling evidence to the prevailing levels of genome wide homozygosity in the subjects. Individuals born from closely related parents often carry genetic burden, with several stretches of DNA inherited from a common ancestor. These sequences characterized by continuous homozygous sites, ROHs, increase the likelihood of expressing deleterious mutations^35^. At the population level, inbreeding decreases genetic variability, with the length of ROH regions reflecting the number of generations since the inbreeding event. Our findings show high frequencies of both short and long ROH tracts, indicating a combination of founder events and high endogamy rates in the groups. These insights have significant implications for assessing the long-term effects of inbreeding on human health and utilizing ROHs to identify loci susceptible to recessive disorders.

We identified clinically significant genetic variants in both known and novel sets across the four populations. Notably, we found a large number of novel exonic variants (1,284) with a minor allele frequency (MAF) > 2%, highlighting the genetic diversity and distinctiveness of Indian populations. These novel variants are unique to their respective populations, likely a result of strict endogamy practices. Twenty three of these novel variants have been annotated through *in silico* analysis as potentially deleterious, impacting protein stability. Their absence in homozygous form suggests that they may be pathogenic recessive variants.

Given the occurrence of founder events, we anticipated a high load of deleterious variants and an increased risk of recessive diseases, similar to observations in Finns and Ashkenazi Jews. We identified several pathogenic/likely pathogenic variants and predicted many potentially pathogenic/loss-of-function (pLoF) exonic variants across the four populations. Most samples were heterozygous for known pathogenic mutations, particularly for rare monogenic disorders, such as mutations in the VWF gene causing autosomal recessive Von Willebrand disease. For some genes, we found that the presence of paralogs allows for greater tolerance to loss-of-function variants^36^, while others are linked to disorders with variable penetrance or are influenced by additional regulatory and external factors, such as NQO1 gene variants^37^, which are key in breast cancer progression. Additionally, several variants showed significant deviations from their typical occurrence rates in South Asian populations.

Apart from the genetic disease risk, understanding diversity in the genetic variants impacting an individual’s response to medications holds crucial clinical implications. There is a global emphasis to increase pharmacogenetic testing to ensure drug safety and increase drug effectiveness. Recent pharmacogenomic research has brought into light marked inter-population disparities in drug metabolism, therapeutic efficacy and safety profiles, highlighting a pressing concern^38^. In our work, we sought to investigate in our study groups, the variability in the prevalence of the VIP pharmacogenomic variants. We reported the presence of *UGT1A1*80+*28,* known for its association with Gilbert syndrome, and *CYP3A5*3* genetic polymorphisms only in the Kalingas. *CYP3A5*3/*3* results in CYP3A5 non-expressor, exhibiting poor clearance of the drug tacrolimus which is used to prevent post-transplantation organ rejection^39^. While the allelic frequency for *CYP3A5*3* in Kalingas stands similar to literature reported values for the south Asian populations, it is crucial to emphasize here that this uniformity does not universally apply, considering the absence of the allele in the other three groups. We observed a likewise scenario wherein the reported *CYP2C19*3* (a premature stop codon in exon 4) levels in both Kallars and Yadavs differs considerably from the stated frequency of 0.08 among South Indians^40^. Other pharmacovariant of importance is the rs2231142 (421C>A) in the *ABCG2* gene, which decreases the ATPase activity of *ABCG2,* increasing rosuvastatin accumulation in the systemic circulation, a drug used for the management of dyslipidemia and coronary heart disease. Second is the rs2108622 in the *CYP4F2* gene linked to altered vitamin K_1_ metabolism, wherein the rs2108622-T carriers require a higher warfarin dose. Furthermore, we also document the presence of rs3918290 and rs2297595 variants in the *DPYD* gene, related to severe toxicities in cancer patients treated with fluoropyrimidines like fluorouracil.

Our study highlights a significant prevalence of Ankylosing Spondylitis (AS) within the Reddy population, primarily linked to the *HLA-B27:04 risk allele.* AS is a highly familial disease, with *HLA-B27* contributing to 20.1% of its heritability^41^. *While data from the Indian subcontinent is limited, HLA-B*27:04 and *HLA-B*27:05* subtypes have been noted as highly prevalent in AS cases in South India^42,43^. To investigate this further, we collaborated with hospitals and found that the KIMs hospital in Hyderabad reported approximately ∼140 patients to be Reddys from a total of 2,963 AS-positive cases. Notably, 28.6% (40 in 140) of these Reddy AS cases were from a particular geographical region, which includes our sample collection area and nearby regions. Further, *ERAP1,* the second most important gene known to be definitely associated with AS, is highly polymorphic and is known to exhibit genetic diversity across different ethnicities. However, the association between *ERAP1* and AS is thought to be attributable to combinations of haplotypes, affecting *ERAP1* function^44^. We discovered four distinct haplotypes not previously reported, despite our small sample size. These patients exhibited typical AS symptoms such as peripheral joint and lower back pain, stooped posture, and skin rashes. Studies on *ERAP1* haplotypes highlight the gene’s role and its interaction with HLA genes in disease, though the prevalence and functional distinctions of specific haplotypes remain debated. The presence of *HLA-B27*, combined with a family history of AS, significantly increases the risk of developing the disease^45^, raising serious clinical concerns for the Reddy population due to the high incidence rate.

In yet another similar example of population-specific disorder, we would like to highlight the high incidence of Epidermolysis Bullosa in Kallar group from an adjacent district to our area of sample collection (unpublished work). However, we do not observe such phenotype in our Kallar samples. Both these examples underscore the prevalence of endogamy in India not only at the population level, but also at the sub-population level, within small geographical regions. Disease associated variants can gain prominence in a founder population with successive generations, more so compounded by inbreeding.

Our study highlights the importance of understanding the clinical and pharmacogenomic impacts of founder events and endogamy in the Indian populations. Population-specific disparities in the field are of high relevance in the context of large-scale genetic inquiries and for nuanced disease risk assessments. Our findings underscore the need for further research to validate and extend these insights. We advocate for creating an accessible database categorizing genetic variations by ethnic background, facilitating precise genome scans for disease risk and drug-response-related polymorphisms. This approach aims to enable tailored treatment strategies, ensuring personalized medications based on unique genetic profiles. This could further help in development of health policies that establish guidelines for specific risk groups. This would mark a pivotal step towards eliminating pathogenic variants and reducing the occurrence of diseases in vulnerable populations.

## Supporting information

Supplementary images

## Data availability

The raw sequence data for the samples included in this study has been submitted under the BioProject ID PRJNA1112977 (to be released on manuscript publication).

## Authors contributions

PM - study design, conceptualization, methodology, sample collection, data generation, data analysis and curation, validation, visualization, writing - original draft, review and editing; AG - sample collection, clinical data curation; YE - sample collection, SCM - supervision of clinical data curation and validation; DTS - supervision, resources and visualization; KT - supervision, study design, conceptualization, validation, writing - review and editing, funding acquisition, project administration, resources and visualization.

## Ethics approval

The project was carried out in accordance with the guidelines set by the Institutional Ethical Committees of the Centre for Cellular and Molecular Biology, Hyderabad, India.

## Consent to participate

Informed written consent was obtained from all the participating volunteers included in the study.

## Acknowledgements

We extend our gratitude to all the participants involved in this study. Our sincere thanks to Akshay Kumar Avvaru and Deepak Kumar Kashyap for critically reviewing the manuscript and for informative discussions. We are grateful to Tulasi Nagabandi from CCMB NGS facility for library preparation and sequencing. We are thankful to Payel Mukherjee for demultiplexing the NGS data.

