## Supplementary images for "Endogamy and high prevalence of deleterious mutations in India: evidence from strong founder events"

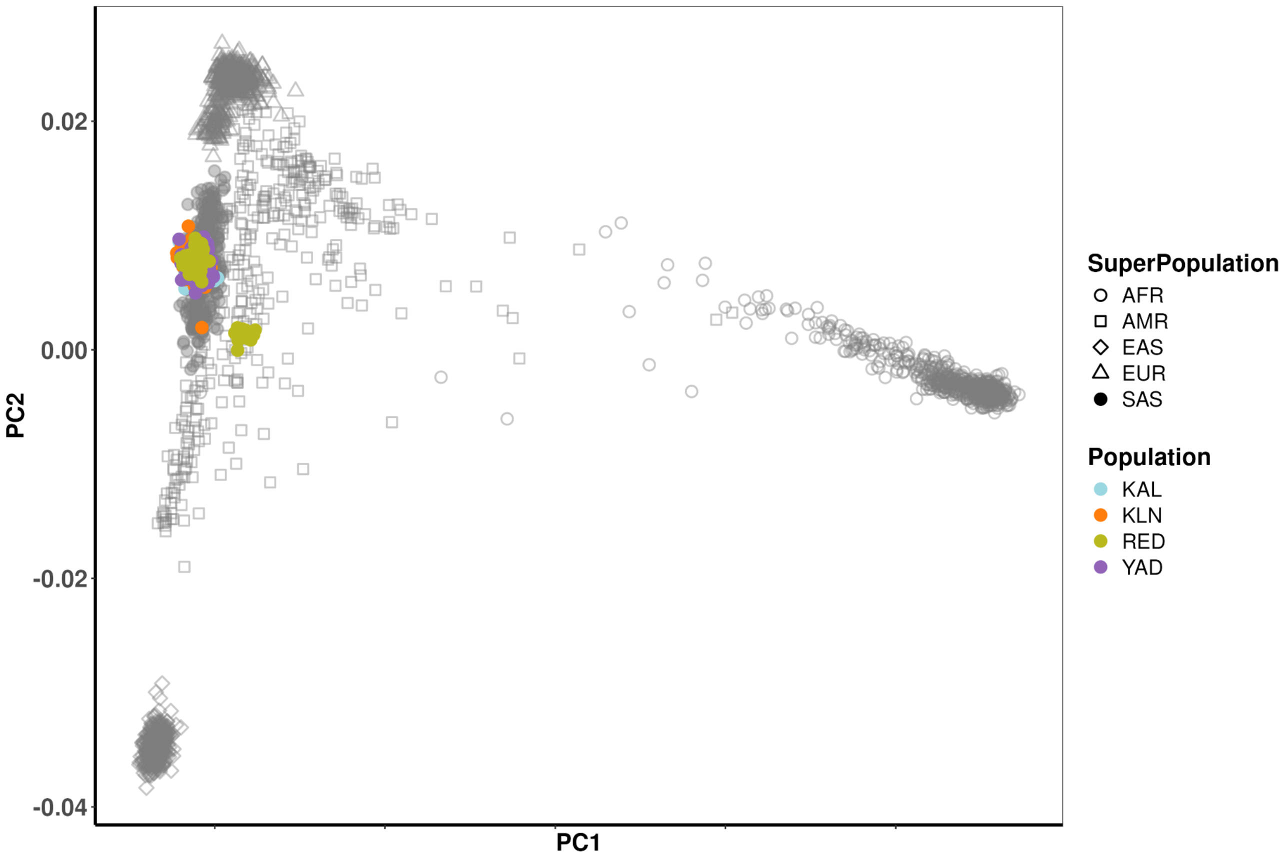


**Fig. 1: PCA plot representing the genetic distance between the four populations and the 1000 Genome dataset, based on their genetic composition.** The four groups cluster with the South Asian populations, showing their genetic proximity.

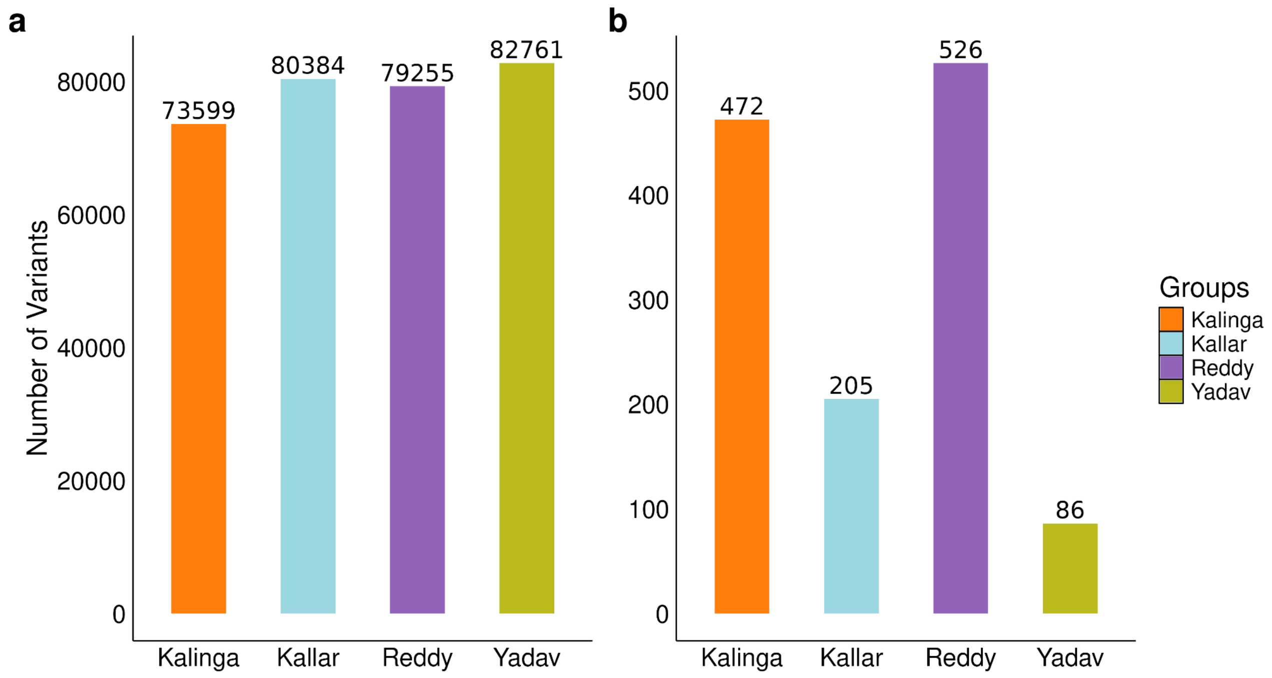


**Fig. 2: The variant stats across the four populations.** (a) Total number of high confidence variants in the exome data (b) Number of novel exonic variants identified in each group in comparison with nine different public datasets. A total of 1,284 unique novel variants were identified across the populations.

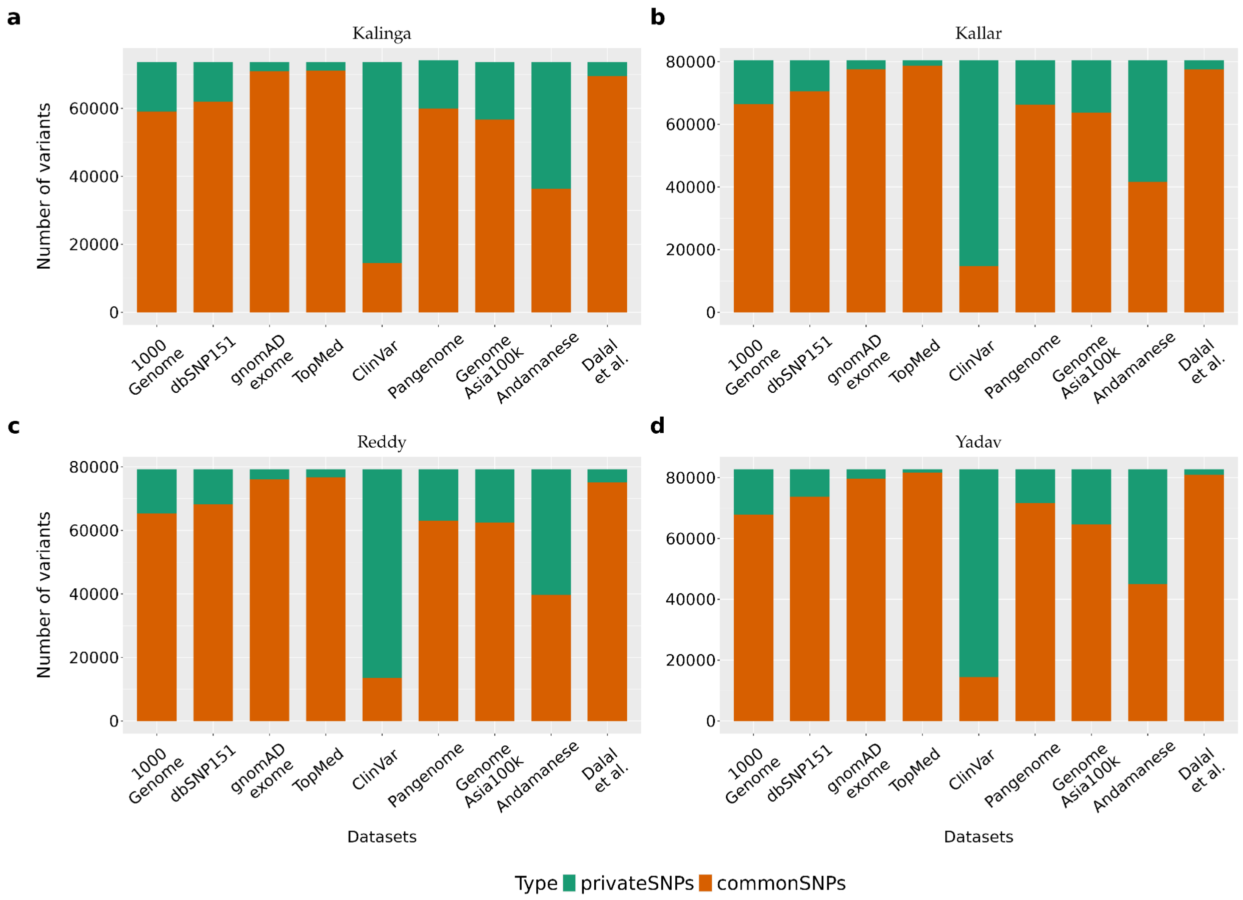


**Fig. 3: Distribution of common and private variants within each group.** The identified variants for each group were compared against different available population datasets. Private variants are the ones that are unique to the corresponding group in contrast to the dataset compared.

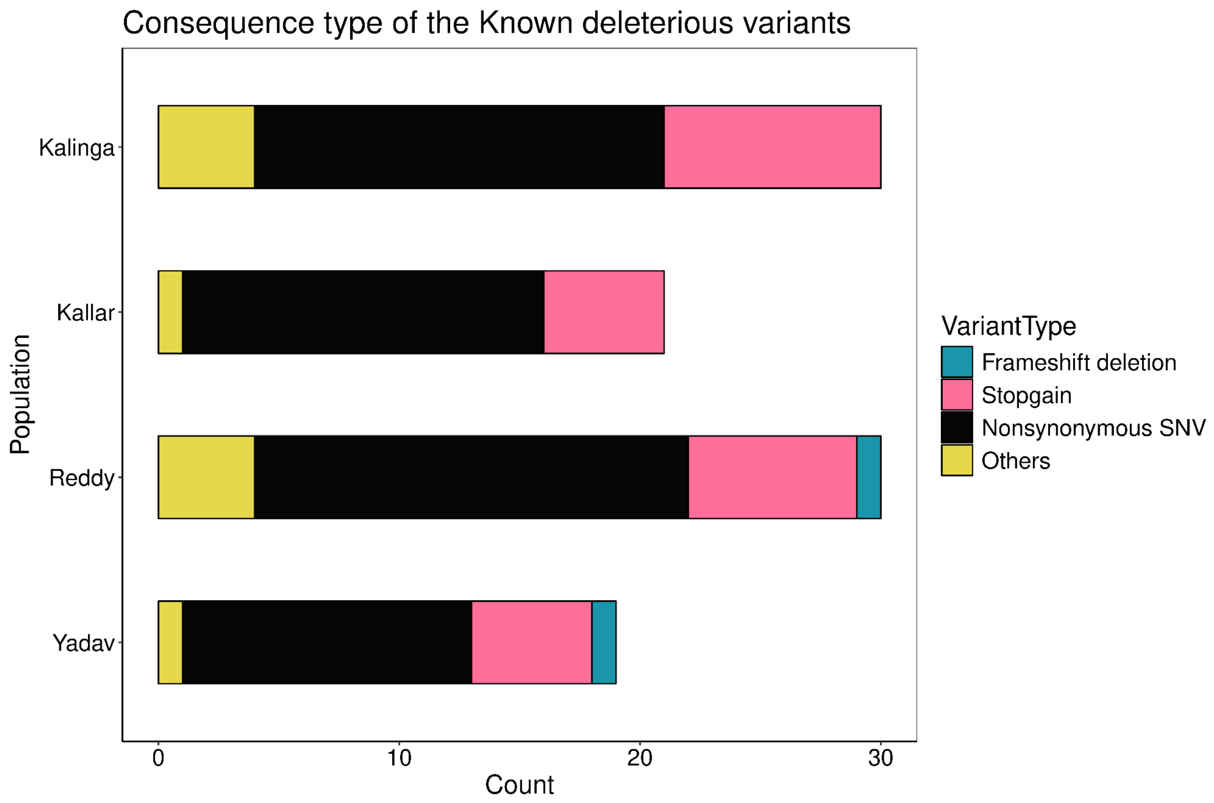


**Fig. 4: Distribution of the consequence type of the known deleterious variants across groups.** Variants annotated to be pathogenic/likely pathogenic in ClinVar are categorized as the known deleterious variants. Majority of them were observed to be nonsynonymous, followed by stop-gain variants.

| Associated.Disorder |
| --- |
| Breast Cancer, poor survival post-chemotherapy |
| Acquired immunodeficiency syndrome |
| Super dismutase, elevated extracellular |
| Delayed puberty, self-limited |
| Pituitary stalk interruption syndrome |
| Von Willebrand disease, type 1 |

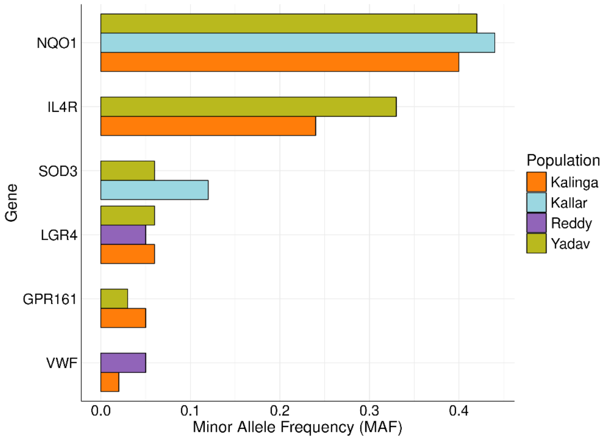


**Fig. 5: Genes with deleterious variants occurring in more than one group.** Six genes were identified to have deleterious variants in two or more groups, with significant AF (p.adj < 0.05). The left panel depicts the disorders associated with mutations in that particular gene. The right panel shows allele frequencies of the variants across the four groups.

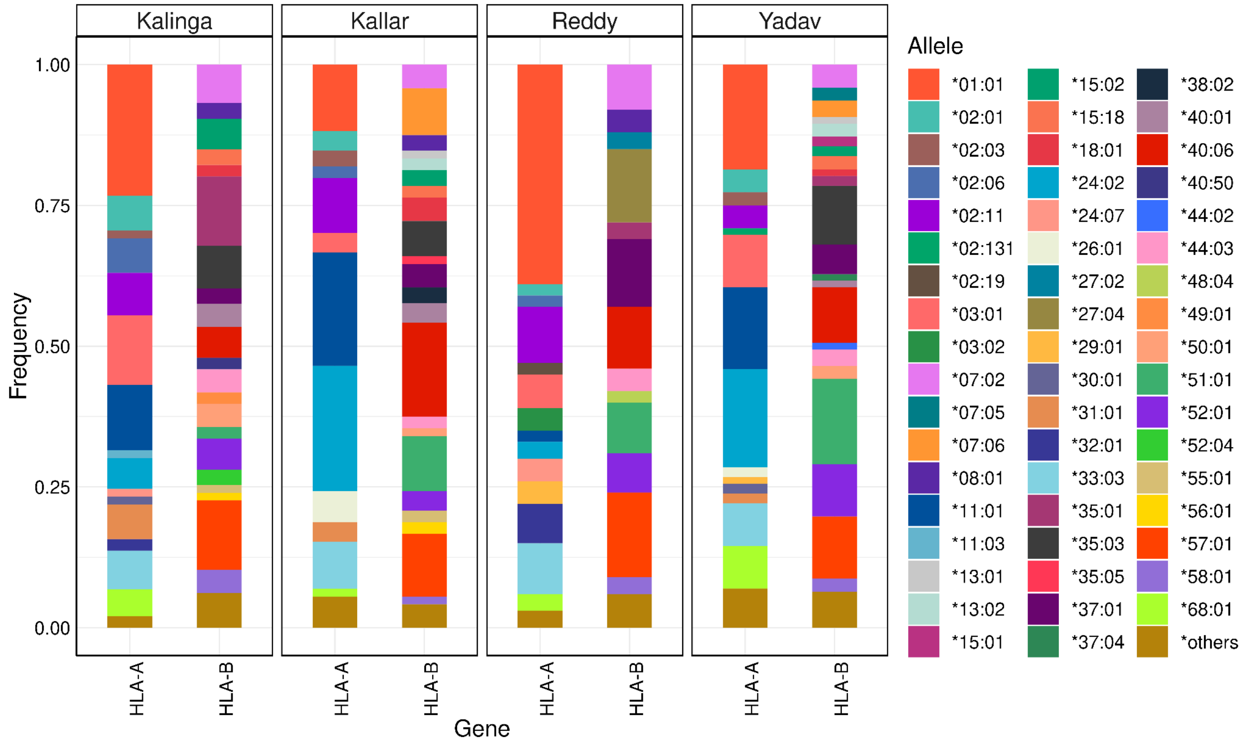


**Fig. 6: Genetic diversity of HLA-A and HLA-B genes across the four populations.** HLA-A and HLA-B are known to be highly polymorphic in nature. The alleles that are harbored by less than 1% of the individuals in a group are clubbed together as others.
